# Investigation of the association between high arachidonic acid synthesis and colorectal polyp incidence: a Mendelian randomisation approach

**DOI:** 10.1101/2022.04.11.22273669

**Authors:** Rachel Moon, J. Bernadette Moore, Mark A. Hull, Michael A. Zulyniak

## Abstract

**Background & Aims:** Arachidonic acid (ARA) is causally associated with colorectal cancer (CRC), a major public health concern. However, it is uncertain if ARA contributes to the development of colorectal polyps which are pre-malignant precursors of CRC. Therefore, this study aimed to investigate the association between lifelong exposure to elevated ARA and colorectal polyp incidence using Mendelian randomisation.

**Methods:** Summary level GWAS data from European, Singaporean, and Chinese cohorts (n=10,171) identified 4 single nucleotide polymorphisms (SNPs) associated with blood ARA levels (p< 5 × 10^−8^). After pruning, 1 SNP was retained (rs174547; p=3.0×10^−971^) for 2-stage Mendelian randomisation to infer the causal effect of ARA on self-reported colorectal polyp outcomes within the UK Biobank (1,391 cases; 462,933 total).

**Results:** No association between ARA and colorectal polyp incidence was observed [OR= 1.00 (95% CI: 0.99, 1.00); P-value = 0.50).

**Conclusions:** Blood levels of ARA do not associate with colorectal polyp incidence. This work supports the contention that downstream lipid mediators, such as PGE_2_, are key for polyp formation during early-stage colorectal carcinogenesis

## Introduction

Colorectal cancer (CRC) is a major global public health concern, accounting for approximately 10% of all global cancer cases. Colorectal carcinogenesis is associated with non-modifiable (e.g., age, ethnicity) and modifiable risk factors such as excess body weight [1]. Diet alone accounts for ∼5% of all CRC cases, with some nutrients more strongly associated with CRC risk[1]. Recently, a Mendelian randomisation (MR) study demonstrated that the long-chain omega-6 polyunsaturated fatty acid (LC n-6 PUFA) arachidonic acid (ARA) is causally associated with risk of CRC (odds ratio (OR) = 1.08 (95% CI 1.05, 1.11 per SD; P = 6.3 × 10^−8^) [2]; however, it is not clear if ARA contributes to early predictors of CRC.

The majority of CRCs are believed to occur via the benign precursor colorectal polyp (conventional adenoma or serrated polyp) in a process that can take approximately 10 years [3]. Therefore, one can hypothesise that ARA exposure is also involved in colorectal polyp development and may offer an early opportunity to mitigate early stages of colorectal carcinogenesis. To date, a single case-control study (n=909 cases, n=855 controls) has applied Mendelian randomisation to examine this and found no association (OR 1.07; 95% CI:0.97–1.02; P=0.41) between a genetic variant within *fatty acid desaturase 1* (*FADS1*; rs174537), which is involved in the ARA synthesis from precursor PUFAs (i.e., linoleic acid), and colorectal adenomas adenoma risk [4, 5]. However, the study reported a required odds ratio of 1.6 to achieve sufficient confidence and minimise the risk of a type II error (i.e., false positive). Given the magnitude of association observed between ARA and CRC (OR = 1.08), it possible that the study was underpowered and that a larger study is needed to test for a casual association of smaller magnitude.

Therefore, to overcome any limitation of power, build on emerging evidence, explore alternative variants, and provide greater certainty regarding the causal role of ARA exposure on colorectal polyp risk, we applied an MR approach in a large prospective UK cohort (n≈500,000) to ascertain if prolonged exposure to elevated ARA synthesis is causally associated with colorectal polyp formation.

## Material and Method

We performed 2-sample Mendelian randomisation using MR-Base[6] with summary data from publicly available GWAS databases. All studies and consortia accessed in the present study on MR-Base were approved by their respective ethics committee, and the subjects from all the cohorts provided written informed consent.

### Sample 1

Single nucleotide polymorphisms (SNPs), associated with ARA at a significance level of p < 5 × 10^−8^, were identified within MR Base. From the Cohorts for Heart and Aging Research in Genomic Epidemiology Consortium (CHARGE; n=8,631 white Europeans; 55% women) and the Singapore Chinese Health Study (SCHS; n= 1540), 6 potential SNPs were identified. The association between ARA and polyps will be investigated in a primarily white European population), which positions SNPs from CHARGE as most suitable. However, to maximise the number of potential IVs we will consider all promising SNPs and perform a stratified cohort analyses if a mixture of SNPs are retained. Of the 6 SNPs identified, two were removed — rs1741 (*PDXDC1*) and rs16829840 (*TMEM39A*) — because their mechanism of association with ARA could not be identified, which risks violating two (i.e., ‘independence’ ‘exclusion restriction’) of the 3 core assumptions of the ‘instrumental variables’ in MR [7, 8]. The 3 core assumptions are as follows: (i) *Relevance*, the variant is associated with the risk factor of interest; (ii) *Independence*, the variant shares no common causes (confounding) with the outcome; and (iii) *Exclusion Restriction*, the variant do not affect the outcome except through the risk factor. Therefore, only rs174547, rs102275, rs174577, and rs174528 were evaluated for inclusion, all of which are associated with ARA synthesis [9].

### Sample 2

The UK Biobank, with self-reported longitudinal data (updated 2018) on colorectal polyps (ukb-b-14210: 1,391 cases of colorectal polyp; 462,933 total), contained all 4 SNPs within its database so proxy SNPs in high linkage disequilibrium (LD; r^2^≥0.80) were not required. Participants were asked to self-report rectal or colon adenoma/polyps via the touchscreen questionnaire. Specifically, they were asked ‘*Has a doctor ever told you that you have any other serious medical conditions*’ and then to select the condition from a panel of options, which included ‘*rectal or colon adenoma/polyps*’. Using MR-Base and the *ld_matrix* function (TwoSampleMR) in R (v.3.5.1), R^2^ > 0.80 were observed between the 4 SNPs within the 1000 Genome project. After pruning for independence (r^2^<0.001), rs174547 was retained as the instrumental variable (IV) for ARA. The rs174547 variant is in very high LD (R^2^ > 0.89) with the rs174537 variant that was tested previously[4], is strongly associated with ARA (β =−1.69 (0.02) % total fatty acids, P-value 3.0×10^−971^), has been used in previous MR studies as an IV for ARA, and demonstrates no association with BMI, smoking, or alcohol intake [2, 5, 6].

### Statistical Methods

The online tool mRnd[10] estimated study power to be over 90% and an F-statistic > 11 to detect a significant difference (P<0.05) in ≥1% change in odds of polyp formation, assuming a conservative mean r^2^ of 0.20 between our instrumental variable and exposure [5, 6]. With a single IV, the Wald estimate was used to evaluate the association between ARA and polyp formation. Brifely, the Wald estimate assumes that the association between the exposure and the outcome (i.e., our association of investigation; β_EO_) is the quotient of the association between IV and the outcome (β_GO_) and the IV and the exposure (β_GE_): β_*EO*_ = β_*GO*_/β_*GE*_. Estimates (β_*EO*_) were exported from MR-Base as logodds and then exponentiated for easier interpretation as odds ratios (ORs), which can be interpreted as odds of reporting one or more colorectal polyps per unit (1%) decrease of ARA (up to most recent reporting period).

## Results

We report a non-significant association [OR= 1.00 (95% CI: 0.99, 1.00); P-value = 0.50] between a 1% reduction of ARA and colorectal polyp risk in the UK Biobank cohort.

In a previous clinical trial, we observed reductions of ARA of ≈1.7% following a 3-month supplementation with fish oil (2 g eicosapentaenoic acid and 1 g docosahexaenoic acid) in young and older men [5]. Therefore, we also present estimates of effect sizes which are proportional to a 1.7% reduction of ARA following fish oil supplementation. The difference in effect sizes between a 1% and a 1.7% reduction of ARA is negligible (log OR_1%_ = 0.000048 versus log OR_1.7%_ = 0.000082) with no change in risk observed following a 1.7% reduction of ARA (OR=1.0; 95% CI: 0.99, 1.00).

## Discussion

We provide greater certainty regarding the association between ARA and colorectal polyp risk in a large UK population. Our results suggest that blood ARA levels are not directly associated with colorectal polyp formation.

The results suggests that despite ARA’s causal association with CRC (OR = 1.08; 95%: CI 1.05–1.11) [2], ARA does not directly contribute to colorectal polyp formation, an early risk-factor of CRC. However, it is plausible that downstream products of ARA (i.e., ARA-derived eicosanoids), rather than ARA-itself, are mediators of colorectal polyp formation and CRC risk (i.e., vertical pleiotropy). This downstream route of investigation is strongly supported by evidence from human and preclinical models that report associations between levels of ARA-metabolising enzymes (cyclooxygenase, COX; lipoxygenase, LOX; and cytochrome P450, CYP), and their products, such as prostaglandin E_2_ (PGE_2_), with colorectal polyp numbers and their transition, and has been recently reviewed [11]. This supports the hypothesis that ARA-derived metabolites, rather than the ARA level itself, promote polyp formation and their transition to malignancy.

We acknowledge two major limitations. First, our use of self-reported polyp incidence is at risk of underreporting in our generally healthy low-risk population (57.4 ± 8.4 years). As the cohort ages and reaches the age of routine polyp screening (i.e., ≥ 60 years), the validly of self-reported data can be tested. Second, our analysis uses summary level data and, therefore, assumes a common effect between randomly assorted exposure groups (i.e., high vs low synthesizers). Future analyses with individual level data with known confounders of ARA metabolism (such as age, BMI, sex, and medication) [1, 5] and colorectal cancer may uncover differences in effect sizes between groups.

In short, this study provides evidence that ARA is not directly associated with colorectal polyp risk and directs future investigations ot focus on products of ARA to appreciate the overarching association between ARA and CRC.

## Supporting information

STROBE Checklist

## Availability of data and material

All data is publicaly available (MR-Base).

## Code availability

MR-Base

## Author Contributions

*RM*: Conceptualization, original draft preparation, formal analyses, writing - review & editing, funding acquisition. *JBM*: Writing - review & editing. *MH*: Writing - review & editing. *MAZ*: Conceptualization, supervision, methodology, writing - review & editing, funding acquisition.

## Ethics approval

All summary level data is publicaly available for use (MR-Base).

## Consent to participate

All summary level data is publicaly available for use (MR-Base).

## Consent for publication

All summary level data is publicaly available for use (MR-Base).

